# Enhancing Rare Disease Education through AI-Driven Podcast Generation

**DOI:** 10.1101/2025.01.27.25320972

**Authors:** Eduardo Perez-Palma, Ilene Miller, Katrine Johannesen, Lauren Chaby, Lindsay Randall, Mike Graglia, Caitlin Grzeskowiak, Lisa Manaster, Laura Lubbers, Amber Freed, Jessica Castrillon Lal, Leah Schust, Terry Jo Bichell, Richard Coca, Victoria Arteaga, Anu Cherukara, Alison Merket, Sebastián Ortiz de la Rosa, Priya Balasubramanian, Ángel Aledo Serrano, Dennis Lal

## Abstract

**Objective:** Rare diseases, including many rare genetic epilepsies and neurodevelopmental disorders, present significant challenges in timely diagnosis, treatment, and patient education due to their rare incidence, complex clinical nature and lack of standardized care pathways. Despite advancements in genetic testing, knowledge dissemination remains inadequate, contributing to delayed diagnosis and inconsistent management. Addressing these gaps requires innovative educational approaches tailored to diverse audiences, including patients, caregivers, and non-specialist healthcare providers.

**Methods:** We explored the potential of AI-driven podcast generation as a scalable solution for rare disease education. We designed a tutorial for the community to use Google’s NotebookLM, a tool powered by Large Language Models and text-to-speech technology to generate podcasts.

**Results:** Eight examples were created from research papers on rare epilepsies (*SCN2A*-, *CACNA1A*-, *SYNGAP1*-, chromosome 8p and *SLC6A1*-related disorders) and a complex genetic research topic (epilepsy polygenic risk scores) in English, Spanish or German. The AI-generated podcasts featured conversational overviews delivered by virtual hosts, with customizable style and tone enabling personalized content creation for different audiences. Feedback from 20 stakeholders, including patient advocacy leaders, researchers, and clinicians, highlighted strong enthusiasm for this approach, particularly for under-resourced patient communities. Respondents praised the accessibility, quality of language translation and educational value of the podcasts, noting their potential to bridge the gap between complex research findings and practical patient care. Key recommendations for improvement included ensuring scientific accuracy through expert review, enhancing clinical depth, reducing redundancy, and incorporating structured episode elements such as introductions and summaries.

**Significance:** The study underscores how AI-driven podcasting can democratize access to high-quality medical information, offering a flexible, multilingual platform that adapts to the needs of global rare disease communities. By refining this approach to include greater oversight and targeted content development, AI-generated educational podcasts could play a pivotal role in rare disease knowledge dissemination, ultimately improving patient outcomes and empowering stakeholders across healthcare systems.

**Key points:**

- AI-driven podcasts bridge knowledge gaps in rare disease education via scalable, multilingual, and accessible formats.
- Our method enables rapid podcast generation, offers tailored content for diverse audiences in various languages with fair clinical depth.
- AI podcasts reduce barriers for caregivers, patients, and clinicians, democratizing access to medical knowledge.
- Human oversight and expert supervision are required and ensures scientific accuracy, balancing accessibility with professional rigor for impactful education.

## Introduction

Rare diseases present unique challenges for patients, caregivers, and healthcare providers due to their complex clinical nature and the limited understanding of their underlying mechanisms.^1^ In the United States, a rare disease is defined as one affecting fewer than 200,000 individuals, yet collectively, rare diseases impact over 30 million Americans^2,3^. Despite advances in genetic testing, the absence of standardized diagnostic pathways and treatment protocols results in prolonged diagnostic delays and inconsistent care. With approximately 80% of rare diseases having a genetic basis, timely access to accurate information is critical for improving patient outcomes^3^.

One of the greatest barriers in rare disease management is the information gap faced by various stakeholders^4^. Caregivers, often functioning as primary decision-makers^5^, need reliable resources to navigate complex care requirements. Also, patients benefit from clear, actionable information to manage their conditions and participate in shared decision-making^6–11^. Meanwhile, general practitioners and non-specialist clinicians frequently encounter rare diseases without adequate exposure or training, further complicating diagnosis and treatment^10^. The limited availability of high-quality, accessible educational resources exacerbates these challenges. Diversified educational formats, such as podcasts, offer an innovative solution by delivering content in a more engaging and widely accessible manner. Research on health-focused podcasts has demonstrated their potential to enhance understanding and retention of complex medical information, particularly among underserved populations.^12–16^

The integration of artificial intelligence (AI) and Large Language Models (LLMs) into education is transforming the way information is delivered^17^, making it highly adaptable to diverse audience needs.^18–20^ For example, Google’s NotebookLM offers features like dynamic “deep dive” discussions and personalized prompts, enabling AI-driven podcast generators to produce engaging, tailored content efficiently.^21–25^ LLMs can process large datasets and generate coherent, contextually rich outputs tailored to specific audiences. By leveraging tools such as AI-driven podcast generators, educators can create scalable, personalized content with minimal human intervention. Incorporating multimedia elements such as slide decks and images into audio overviews further broadens the scope of content delivery, making the process versatile and impactful. Text-to-speech technology further enhances this process, enabling the rapid transformation of text into high-quality audio content. These AI-driven solutions not only reduce production time but also allow customization in tone, complexity, and style to cater to diverse listeners.^18,20,22–26^

This study examines the potential of AI-driven podcast generation to improve rare disease education. We use five rare genetic epilepsies, neurodevelopmental disorders, and a complex genetic research topic as case studies, presented in English, Spanish, and German. By automating the creation of accessible and engaging audio content, this approach seeks to bridge critical gaps in rare disease knowledge. Emphasizing scalability, personalization, and adaptability, the study demonstrates how AI can democratize access to high-quality educational resources, ultimately empowering patients, caregivers, and healthcare professionals.

## Materials & Methods

From list of papers to podcast. NotebookLM served as the primary tool for podcast generation. It is an AI-driven research assistant capable of processing user-uploaded multimodal content, including research papers, slide decks, and images, to generate accurate, contextually grounded outputs. Its source-grounded approach ensures that all content generated is directly derived from the uploaded materials, promoting accuracy and traceability.^21,22,24,25^ Audio Overview Feature: A core feature of NotebookLM used in this study was the “Audio Overview,” which transforms uploaded content into a podcast format. This feature generates a dynamic “Deep Dive” discussion between two virtual hosts, providing a comprehensive and conversational overview of the selected topic. The addition of multimedia inputs enhances engagement and provides users with a richer educational experience by allowing AI to infer meaning from visual data. The output is generated using text-to-speech technology, allowing users to listen at adjustable playback speeds. Large Language Model: At the heart of NotebookLM is a sophisticated LLM that processes the uploaded documents, extracts key insights, and generates scripts for podcast production. The user can customize the podcast’s style, tone, and complexity to suit different audiences. Currently, the NotebookLM supports the upload of up to 50 files per notebook, with a maximum file size of 200 megabytes or 500,000 words, enabling comprehensive coverage of a given topic.^24^

## Results

### 1. Tutorial

#### Step 1: Selection of Relevant Research Papers

Research papers related to for each case study were identified through pubmed, google scholar or based on domain expertise by the authors. Selection criteria included scientific rigor, relevance to rare disease education, and the potential to provide valuable insights for non-specialist audiences. Both original research articles and review papers were included to ensure balanced content.

#### Step 2: Uploading Papers and Generating Structured Prompts

The selected research papers were uploaded to NotebookLM in PDF format. Once uploaded, the tool automatically processed the documents, generating summaries, key facts, and a knowledge base for podcast creation. The “Audio Overview” feature was then used to initiate podcast generation without requiring additional user prompts.

#### Step 3: Special Instructions for Language-Specific or Topic-Refined Episodes

The podcast can be further customized by including special instructions to tailor it for a specific language-speaking audience or topic to focus on. For example, when creating the German-exclusive episode, the following prompt was used where “German” or ‘Spanish’ was added for “[Language]”:

#### Example prompt for special Instructions

- This episode will only be in [Language]. All discussions, interviews, and commentary must be conducted in [Language] for the entire duration of the episode.
- No English or other languages should be used in the conversation, except when absolutely necessary to clarify a term or concept unique to a specific language.
- Future translation into English is planned, but for now, the episode is [Language]-exclusive.

This type of prompt ensures that the generated content is fully adapted for non-English-speaking audiences, broadening accessibility and reach.

Step 3: Generating and Reviewing the Audio Overview

The “Audio Overview” feature produced a podcast with two virtual hosts engaging in a scripted discussion. The initial audio output was reviewed to ensure factual accuracy, clarity, and overall quality. NotebookLM’s automatic citation generation facilitated the verification of all content against the original sources.

## 2. Case studies

Given the diverse target audience across rare epilepsies—including non-expert caregivers and patients—ensuring content clarity and accessibility was a primary focus. Research papers and reviews that could be effectively summarized and presented in a conversational style were prioritized to enhance comprehension and empower listeners in managing rare diseases. The tutorial outlined a step-by-step process for generating AI-driven educational podcasts, resulting in eight case studies that cover rare disorders and complex topics in either English, Spanish or German. Below are examples of AI-generated podcast content in English and other languages.

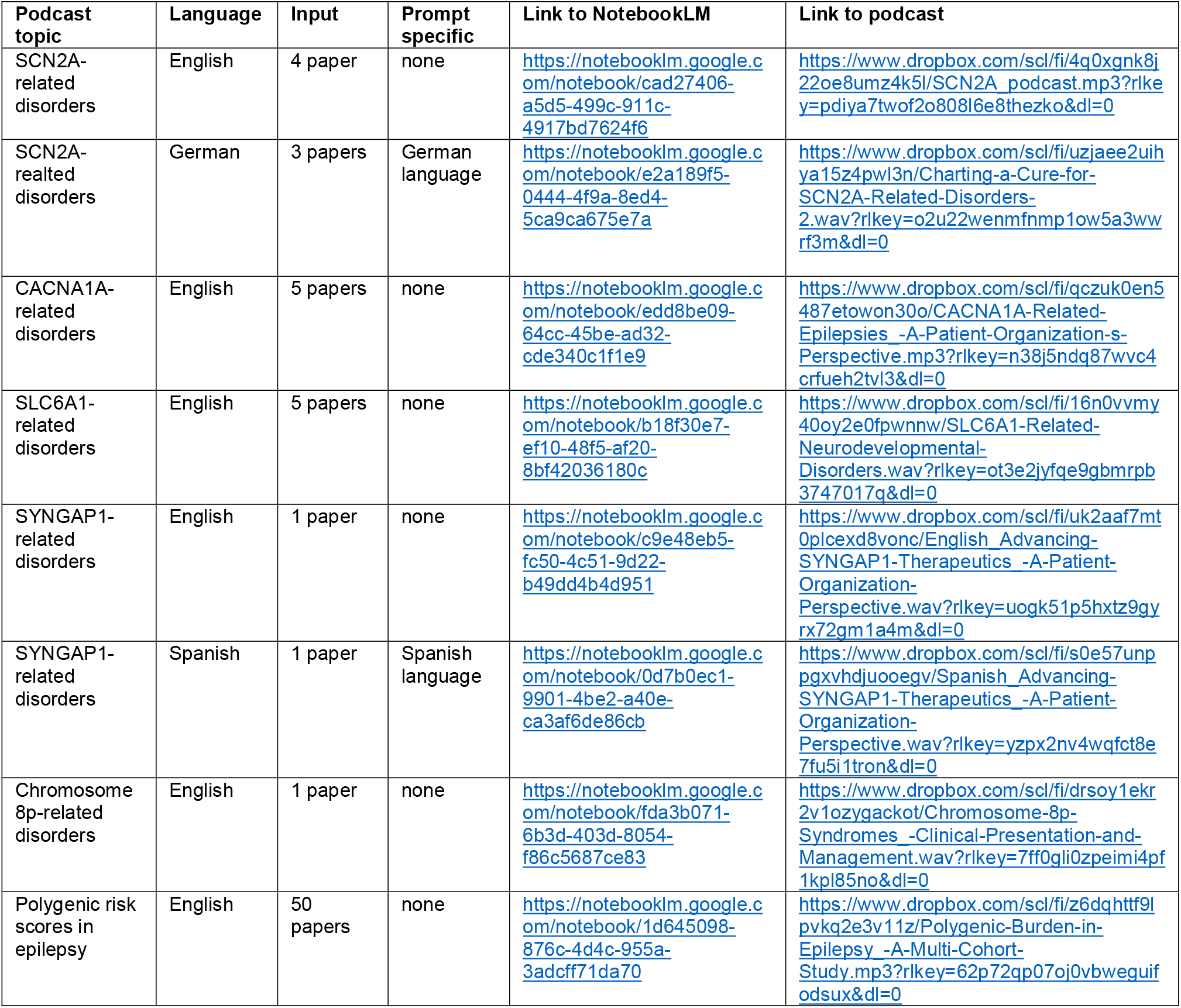

## 3. Expert Feedback on AI-Generated Podcasts for Rare Epilepsy and Neurodevelopmental Disorders

To assess the feasibility and acceptability of AI-generated podcasts as an educational tool for rare epilepsies and related neurodevelopmental disorders, feedback was gathered from 20 key stakeholders representing a broad spectrum of expertise. This multidisciplinary group included leaders of patient advocacy organizations, clinical researchers in rare epilepsies, and basic scientists specializing in genetics and pharmacology. Their responses underscored the potential benefits of this approach while highlighting opportunities to enhance accuracy, consistency, and depth of content. Full responses are available in the supporting information document.

### 3.1. Overall Acceptability and Educational Value

Across multiple respondents, there was strong endorsement for the use of AI-driven podcasts in the dissemination of complex medical information, particularly for under-resourced patient communities. Several feedback sources emphasized that these podcasts could reduce barriers to understanding rare diseases, enabling caregivers, clinicians, and non-specialists to grasp essential information more efficiently: “This could be an incredible resource for rare epilepsies— especially those that are ultra-rare and under-resourced.” (Feedback 1, see Supp. Data) Similarly, stakeholders mentioned the ability of AI-generated audio content to accommodate time-constrained audiences, such as families and healthcare providers: “As this technology matures, I imagine this will have enormous value especially for non-scientists who may struggle reading scientific papers.” (Feedback 5, see Supp. Data) By presenting scientific findings in clear, conversational formats, AI-generated podcasts were perceived as a promising avenue for bridging the gap between research publications and day-to-day clinical practice in epilepsy and neurodevelopmental disorders.

### 3.2. Accuracy and Content Validation

While enthusiasm for the podcasts was high, several respondents highlighted the need for rigorous quality control and domain expertise in their development. For instance, the potential for oversimplification of complex topics—such as antiepileptic mechanisms or advanced genetic therapies—was noted: “Some of the things that are stated are a little bit oversimplified … 4PB appears to be a drug for SLC6A1 only.” (Feedback 2, see Supp. Data) To address this concern, reviewers advocated for structured verification: “Endorse your suggestion of having a lay/PAG [patient advocacy group] and clinical domain expert as a reviewer for accuracy/reliability.” (Feedback 1, see Supp. Data) Respondents also mentioned terminology inconsistencies and mispronunciations, particularly with regard to key proteins and pathogenic variants (e.g., “GAT1,” “disease-causing variants” instead of “mutations”). Such issues underscored the importance of leveraging expert input and editorial oversight to maintain scientific rigor and clarity.

### 3.3. Depth of Content and Structural Clarity

Several respondents welcomed the podcasts’ conversational approach but recommended greater detail in discussing clinical features, seizure phenotypes, and management guidelines: “I would have liked more discussion about the symptoms and natural history of the disease … what can families expect along the continuum?” (Feedback 1, see Supp. Data). Reviewers additionally encouraged the incorporation of more real-world, practical guidance, particularly in navigating healthcare systems, insurance, and genetic testing referrals. Feedback highlighted the importance of integrating detailed clinical insights and practical recommendations to enrich the educational value of the podcasts.” (*“Scientific metaphors and emphasis on key points make it easy to listen to”* - Feedback 15). Beyond clinical detail, multiple participants requested explicit mention of relevant websites, foundations, and advocacy organizations, ensuring that patients and families could access further resources. A recurring observation was the tendency for certain segments to repeat or revisit the same topics. Ensuring that episodes are carefully edited to minimize redundancy and to follow a clear thematic structure was deemed essential to maintain engagement: “Each topic is only covered very briefly … it could have been nice to dwell a little bit longer and share more details.” (Feedback 2) “There is some circularity in the podcast, where the same topics are touched upon after a while.” (Feedback 2, see Supp. Data).

### 3.4. Tone, Format, and Persona Assignments

The majority of respondents found the two-host format engaging, particularly when the tone replicated a news-studio discussion. However, some emphasized the need to calibrate the degree of informality so as not to trivialize important medical content: “At points the response from the hosts is not completely aligned with what has been said … there is a fine line between accessible language and overly casual in relation to the subject matter.” (Feedback 4, see Supp. Data). Overall, the two-host format was praised for its relatability, but listeners suggested refining the roles to ensure consistency and avoid tonal shifts (*“The hosts did a good job at guiding the conversation but could improve consistency in their roles with better prompting”* - Feedback 17). Here, inconsistencies in host knowledge level were also cited, with one “host” occasionally posing basic questions and later providing high-level expert responses. A clear delineation of the host personas—such as an “expert” and a “learner”—was recommended to maintain a cohesive and credible narrative (Feedback 12, see Supp. Data). Additionally, reviewers suggested adding a brief introduction, objectives, and closing statements in each episode to offer listeners a structured roadmap.

### 3.5. Potential for Customization and Multilingual Outreach

Stakeholders highlighted the successful production of episodes in languages other than English, demonstrating broader international applicability. This feature was particularly valued given the multinational nature of rare epilepsy communities. Tailoring content for multilingual and diverse audiences allows for broader reach and inclusivity in global rare disease education. (*“Spanish podcasts offer an opportunity to share essential updates in rapidly evolving fields”* - Feedback 18). Moreover, the capacity of LLM-powered tools to generate custom content for specific groups (e.g., clinicians vs. caregivers) was noted as a potential advantage: “With the right prompts you can make sure that the information you want to relay is included … and provided in an understandable way.” (Feedback 2, see Supp. Data) Coupled with user-friendly prompt engineering, AI-driven podcast generation opens opportunities for rapid updates, corrections, and expansions to address emerging research or newly approved therapies in rare epilepsies.

### 3.6. Enthusiasm for Future Implementation

Nearly all respondents expressed a desire to leverage the technology and recommended broader distribution and collaborative development. Leaders of patient advocacy groups, in particular, showcased interest in using these AI-generated podcast episodes to disseminate up-to-date scientific findings to their networks: “Wow! I just listened and am blown away … Please share with us how we can share it to the community.” (Feedback 10, see Supp. Data) Several also called for additional tutorials and training, emphasizing the importance of capacity building and knowledge transfer: “UNBELIEVABLE!!! Please teach me.” (Feedback 7). Stakeholders emphasized the transformative potential of AI-driven podcasts for rare disease education, citing their ability to empower communities and foster awareness. (*“This tool has the potential to revolutionize how we address the knowledge and implementation gap in rapidly evolving fields”* - Feedback 16). Their accessibility and adaptability make them ideal for both professional development and community engagement.” (*“This is a great way to break down complex topics into digestible and approachable pieces”* - Feedback 19). Such engagement underscores the collective recognition that AI-generated podcasts, if refined to ensure accuracy, depth, and clarity, have the potential to revolutionize knowledge dissemination in rare epilepsies and neurodevelopmental disorders.

## Discussion

For rare epilepsies clinical and research landscape is rapidly evolving and characterized by diverse and often isolated patient populations^27^. This study highlights the potential of AI-driven podcast generation for addressing critical educational gaps in rare disease management, particularly within epilepsy and neurodevelopmental disorders. By leveraging Google’s NotebookLM to create tailored audio content, we illustrate how complex scientific information can be transformed into more accessible formats for both medical professionals and laypersons. Podcasts offer advantages such as multitasking convenience, personalization to accommodate diverse learning styles, and linguistic flexibility.^18,20^ This adaptability, coupled with near-real-time updates in fields like rare disease research, may significantly broaden the reach of accurate and current medical information.

Although we could generate eight podcasts on epilepsy research papers fast using NotebookLM, multiple content-related challenges became evident based on end-user feedback. Foremost among these is the need for rigorous quality control to maintain scientific integrity and avoid overgeneralization.^24^ In the context of highly specialized or novel therapies, several respondents cautioned against conflating research-stage interventions with approved treatment modalities, underscoring the importance of distinguishing between presenting findings from research vs. indirectly providing (false) treatment recommendations. This extends to ensuring that discussions of investigative drugs—such as antisense oligonucleotides or small-molecule agents—do not inadvertently appear as commercial endorsements. Equally pivotal is the balanced integration of clinical depth and real-world relevance. A number of stakeholders advocated for more detailed coverage of fundamental disease features—e.g., phenotypes, prognoses, and the spectrum of seizure types—so that families and clinicians alike can better contextualize the information within everyday care. Similar issues of accuracy, quality control, and the potential for misinterpretation have been documented in other medical AI podcasts. While prior studies show that podcasts can enhance medical knowledge, they stress the importance of human oversight for credibility. Moreover, AI-generated podcasts face an inherent challenge: oversimplifying complex medical topics. Therefore, striking a balance between relatability and professional rigor remains key to ensuring content is both comprehensible and trustworthy.^18,28–30^

From a listener experience perspective, the dual-host or “ping-pong” format was generally well-received for its approachable style, though issues with tone inconsistency, abrupt shifts in content complexity, and uneven responsiveness were noted. Users suggested assigning distinct roles, such as expert and learner, to enhance clarity and conversational flow. Clear structure— incorporating defined introductions, smooth transitions, and concise summaries—was highlighted as essential for improving coherence and reducing redundancy. Additionally, the importance of expert review (e.g., patient advocacy groups or scientists) was emphasized, not only to ensure accuracy but also to increase the relevance of patient-centric topics and address nuanced language issues. These observations echo findings from previous studies, which similarly point to challenges with tone variability, structural weaknesses, and content accuracy in AI-generated podcasts.^18^ Prior research underscores the importance of employing clear outlines, structured episode segments, and stakeholder engagement to enhance both listener engagement and content credibility^16^ – all currently challenging using NotebookLM, given its limited prompting ability.

Our preliminary experiences with using NotebookLM to transform rare disease manuscripts into podcasts align with multiple reports emphasizing the potential of AI-powered podcast creation to substantially reduce production time—some estimate up to a 70% decrease—while also underscoring the need for vigilant oversight.^25,31^ In our application, this efficiency enabled rapid content generation, which is particularly pertinent given the urgent need to advance knowledge in rare epilepsies, where delayed access to information can significantly impact clinical outcomes^27^. Similar to previous findings^25^, the generative tool demonstrated an impressive ability to synthesize information from various file formats and tailor its focus, yet limitations remained evident in scripted-sounding delivery, restricted customization features, and the risk of factual “hallucinations.” Moreover, requests for greater voice diversity, refined editorial control, and enhanced cultural nuance echo concerns voiced in the broader literature, indicating that although these AI-generated podcasts can dramatically streamline knowledge sharing, they require careful human validation to ensure both scientific accuracy and a tone suitable for diverse patient populations^22,32^. These concerns are critical when addressing conditions like epilepsy, where accurate, nuanced information about seizure types, comorbidities, and emerging treatments is essential^27^. As the technology evolves—especially in expanding multilingual capabilities and refining medical content precision—AI-generated podcasts may become powerful tools for disseminating epilepsy research findings and fostering global collaboration. To realize this potential, ongoing editorial stewardship and close collaboration with epilepsy advocacy groups and clinicians will be key in ensuring that such content is both scientifically accurate and culturally sensitive.

## Conclusion

Taken together, our findings underscore the importance of combining AI innovation with human oversight to produce engaging and scientifically robust podcast content. Our stakeholders have noted the potential for widespread dissemination, particularly in addressing critical educational gaps for rare epilepsy, where timely and accessible information is vital. Future efforts would benefit from systematic evaluation of learning outcomes, with a focus on epilepsy-specific metrics such as seizure management understanding, comorbidity awareness, and treatment adherence. Additionally, direct comparisons with more traditional patient education tools, and best-practice guidelines for ethically responsible AI integration, will be essential. By addressing the limitations identified here—content oversimplification, the need for clinical depth, authenticity in presentation, and conscientious referencing—AI-driven podcasting could become a highly effective platform for epilepsy education and advocacy, empowering both patients and healthcare providers. Ultimately, this approach could bridge gaps in knowledge and support within the global epilepsy community.

## Supporting information

Feedback 5

## Data Availability

All data produced in the present work are contained in the manuscript

