## Supplementary material for "Enhancing Rare Disease Education through AI-Driven Podcast Generation": Supp. Data

**Feedback 1:**

Ilene Miller, Director Rare Epilepsy Network:

“*This could be an incredible resource for rare epilepsies - especially those that are ultra rare, under resourced. Envisioning the impact of one for all 150 rare epilepsy organizations that are part of the Rare Epilepsy network - that would be incredible. Some inputs follow which I hope are helpful - and take or leave as useful -*

- *Endorse your suggestion of having a lay/PAG and clinical domain expert as a reviewer for accuracy/reliability.*
- *Would include references and resources at the end of each episode*
- *Really liked the inclusion of 2 voices (male and female) and the easy back and forth between them - it was very "listenable."*
- *I am curious about the use of the language "genetic medicine" vs. precision medicine related to the discussion of ASOs.*
- *I believe Praxis was mentioned by name. I like the inclusion of a novel therapy in development but suggest thinking through implications of commercial recommendations and whether they will be perceived as endorsements by the the PAG and/or reviewers.*
- *I liked the time frame discussion of setting realistic expectations for ASOs and gene editing - few years and longer - but would want to be sure that is accurate to create hope but not false expectations for families*
- *The discussion about the importance of the "ripple effect" of a deep dive into one gene was a good point but including a specific example or two of that would be more compelling; this felt a bit empty/light*
- *I liked the personal stories as a way to elucidate the everyday challenges but would have liked more discussion about the symptoms and natural history of this disease - what can families expect along the continuum; what are the breadth of symptoms families are experiencing*
- *I liked the discussion of early interventions beyond medicine including SL, OT, PT - are there others that families might not be aware of? As a parent, I want to know what I can do and when I should do it*
- *In regard to overcoming the challenges of navigating health care - is there more practical advice that can be offered - specific resources in addition to the disorder orgs - financial aid, understanding insurance, accessing genetic testing, getting waivers etc.*
- *Perhaps some missed opportunities for caregiver education include: the importance of getting a referral to a higher level center/specialist, SUDEP and other causes of mortality, seizure action planning,*
- *I don't recall hearing much about the type of seizures that show up and perhaps I missed it but the notion that they can be refractory and the limitations of medicine would be important to explain*
- *Would love to add a little depth and connection to the complex of rare epilepsies - SCN2as is part of a complex of disorders that are part of rare epilepsies, epilepsies, and other buckets to which they are connected*
- *Would love to see inclusion of other resources like REN's website for listeners who may want to learn about other disorders*
- *Finally is the intention to do these in partnership with a single organization and any considerations where  there are multiple organizations for a single disease.*”

**Feedback 2:**

Katrine Johannesen, clinical-genetic researcher on rare epilepsies, Danish Epilepsy Center, Denmark:

“*The language is clear and comprehensible and the ping-pong between the two “hosts” works well, it feels a bit like being in a news studio. Content wise, I feel that some of the things that are stated are a little bit oversimplified, eg. 4PB appears to be a drug for SLC6A1 only, and ravicti is described as a drug, where 4PB is part of it. I also feel that for educational purposes the podcast is missing some basic information about SLC6A1-related disorders, such as phenotype, prognosis, and recommended treatment, which why by information that clinicians and families would ask for. Still, the podcast covers some of the important topics concerning SLC6A1-related disorders including 4PB and clinical trials, including genetherapy.*

*While the conversation between the two hosts works well, at around 5.30 minutes into the podcast, the topics begin to repeat themselves. Further, each topic is only covered very briefly, where it could have been nice to dwell a little bit longer with each topic and share more details.*

*I really like the part on Amber Freed and SLC6A1 and what they have done for the community.*

*Minor comments would be to adjust language, eg. Pronounciation of GAT1, not spelled out, and using disease-causing variants instead of mutations.*

*I guess most of the issues outline above can be corrected through prompting.*

*I also listened to the SCN2A podcast: The language is clear and comprehensible and the ping-pong between the two “hosts” works well, it feels a bit like being in a news studio.*

*The introduction to Nav1.2 was good, and also the explanation of GOF vs LOF and the phenotypic spectrum correlated to functional effect. Again, love that the SCN2A foundation is included in the podcast. For families I think this is important, knowing and having explained in an understandable language, what the patient foundation provides.*

*ASOs is also explained in a really nice way, and the amount of time used to explain this is appropriate, the topic is explained in detail, and not just rushed by.*

*I think something is wrong at around 15 min, where the hosts say goodbye, but then it seems like the podcast rewinds to a previous section?*

*Then I listened to the polygenic risk score because I was curious and have not read the papers myself: I really like the way it seems that the hosts have a conversation, even with like little pauses. Makes it very real.*

*Very relevant discussion on epilepsy severity and PRS score.*

*Also, very nice that it is highlighted that correlation is not equal to causality, I believe this is important when targeting a non-expert audience. And the same when it is highlighted that PRS scores should not just be handed out as reports but should be provided in the context of genetic counselling.*

*Again, I’m missing an explanation on what PRS scores are, and what the scores are derived from. But the section on what you can use a PRS score to after having your first seizure is good.*

*I really like the section on what patients could use polygenic risk scores too, such as higher awareness in patient with autism with a high PRS for epilepsy. And also, the idea of each of these additional scores or biomarkers, all add up to more personalized treatment.*

*There is some circularity in the podcast, where the same topics are touched upon after a while.*

*All in all, these podcasts hold great promise to provide easy and fast access to relevant information for clinicians and families worldwide, in the relevant language. Of course, the podcasts are not perfect, but they provide a fast overview, and with the right prompts you can make sure that the information you want to relay is included in the podcast and provided in an understandable way. Furthermore, these AI tools make it possible to provide information tailored to eg clinician or families or whoever your chosen audience is.*”

**Feedback 3:**

Amber Freed, founder of the SLC6A1 Connect:

“*I found the podcast to be a powerful tool to disseminate information to a wide variety of audiences.  This would be a great solution for the SLC6A1 community to understand topics in bite sized increments.*”

**Feedback 4:**

Lindsay Randall, CEO of SLC6A1 Connect UK-AQ:

“*It would be interesting to understand the potential for tweaking imperfections that are heard, for example in the SLC6A1 podcast, the term GAT-1 is communicated in several different ways throughout, and there is also some repetition in content. At points the response from the hosts is not completely aligned with what has been said, and that can make the discussion feels disingenuous, and there feels a fine line between the hosts language being accessible, and overly casual in relation to the subject matter.*

*Overall this idea feels like a tremendous opportunity in terms of presenting complicated information in a more accessible and understandable way, particularly for patients, caregivers, and even professionals who are just learning about a new condition, but caution and scrutiny over the final product and the way the hosts present the content is important. I am excited to hear more from this project and SLC6A1 podcasts to share with our patient community, and interested professionals.”*

**Feedback 5:**

Alfred George, Professor and Chair of Pharmacology Northwestern University:

*“This is very impressive! As this technology matures, I imagine this will have enormous value especially for non-scientists who may struggle reading scientific papers.”*

**Feedback 6:**

Caitlin Grzeskowiak, PhD, Chief Research Officer at Epilepsy Foundation:

*“I would be very interested to know how the structure of these podcasts were prompted- there are definitely a few areas (I listened to SCN2A podcast) where I could tell the info was scrubbed from site (question around 3:13 minute mark) but definitely with a human verification this has incredible potential and would give average listeners/newcomers a very digestible source of info. Thanks for sharing this!! Would love to see the tutorial and comment.”*

**Feedback 7:**

Terry Jo Bichell, Director of Combined Brain:

*“UNBELIEVABLE!!! Please teach me.”*

**Feedback 8:**

Laura Lubbers, Chief Scientific Officer at Cure Epilepsy Foundation:

*“Listened to the one on PRS and I have to say, it’s pretty amazing. I’m going to share it with our Comms group for more feedback. Thanks so much for sharing this!”*

**Feedback 9:**

Priya Balasubramanian, Director of Research. Staff, Board and Advisory, CURE Epilepsy

*“Thanks so much for the opportunity to comment on the paper. I think this could be such a powerful tool in our field enhancing education opportunities for advocates, scientists, and clinicians.”*

**Feedback 10:**

Leah Schust, president of the FamlieSCN2A Foundation:

*“Wow! I just listened and am blown away!  Please share with us how we can share it to the community! Love it.”*

**Feedback 11:**

Lisa Manaster, Co-Founder and President of the CACNA1A Foundation:

*“This is hysterical! How did you do this? Has anyone ever listened to the podcast Pivot with Kara Swisher and Scott Galloway? The speakers sound just like them!  "on the verge of a new era..." exciting! “*

**Feedback 12:**

Eduardo Perez-Palma, Assistant Professor, Universidad del Desarrollo, Chile:

*“Overall is an impressive output in such a short period of time. if I was a student I would definitely use this strategy to learn when I am on the move or doing dishes. Assuming the content and redundancy can be edited (as Katrine pointed out), my main comment is: that at some moments the podcast felt inconsistent when the same character would ask very basic, almost introductory questions at one point and then later in the conversation provide detailed, expert-level answers or comments. This inconsistency disrupted the flow and made the dialogue feel less natural, as it’s not typical for a single person to switch between such extremes of knowledge within a conversation. It might help to assign distinct personas to the speakers, with clear roles—for example, one as the expert and the other as the learner—to create a more intuitive and engaging listening experience.”*

**Feedback 13:**

Lauren Chaby, Project 8p Foundation:

*“Such a cool idea! it's a nice question and answer style that is more engaging and would make publications more accessible to a wider audience, which is of high value,*

*overall, it's really neat, with some opportunities for improvement, here are some more specific thoughts:*

- *the pauses and transition between "speakers" seem artificial at times*
- *it's unusual to not have any kind of intro, it jumps straight into discussion*
- *mispronunciations including indupdel*
- *it's typical to do some kind of intro of the authors / studies, which wasn't done, so it's not clear who did the study or why, or how often these types of clinical recommendations are done for rare disease*
- *tone was a bit strange "wow that's a significant finding" was deadpan*
- *non-urbal vs non-verbal, incorrect*
- *there are a couple of spots that have strange noises, I'm not sure if they're supposed to be filler noises but they are a bit odd*
- *a driving "why" was missing that would be part of a typical podcast*
- *recommendations miss the temporal component of recommendations that would be key to interpreting (ie when should I get an MRI / cardiac test)*
- *beautiful messaging of hope and resilience, great messaging to reach out to clinicians and Project 8p, and knowing you're not alone”*

**Feedback 14:**

Mike Graglia, Co-Founder and Managing Director Syngap Research Found

*“Podcasts are a powerful way to bring information to patients and caregivers — SRF has developed three pods to engage and educate our community.  This work, using AI to bring papers  to patients via this medium is desperately needed.  We had 52 papers on SYNGAP1 last year, few parents read them all, but most parents listed multiple podcasts.  This effort can help engage patient caregivers which is requisite for the clinical trials that are imminent.“*

**Feedback 15:**

Sebastián Ortiz de la Rosa, Paediatric Neurologist, Young Epilepsy Section - ILAE

“*I have listened to the PRS podcast, and here are my comments.*”

- “*I find it very helpful how the podcast will help to reduce the knowledge gap and accessibility both for clinicians and patients and for understanding both basic and advanced topics in epilepsy and genetics.*
- *It is extremely important to Involve plain language and patient perspectives in the discussion; this will enhance the engagement of advocates, patients, and families of those affected by genetic disorders.*
- *Prosody, pauses, and emphasis on key points make it very easy to listen to.*
- *The implications of these kinds of tools are yet to reach a ceiling; it is clear that scientific communication is evolving, and plain language, a relaxed environment, stressing the important findings, and using a well-versed base will become a game-changer for the whole community*.”

**Feedback 16:**

Ángel Aledo Serrano, Neurologist and Epileptologist - Director of Vithas Clinical Neuroscience Institute.

“*I listened to both episodes about SYNGAP1. This is my feedback*:”

- “*The quality of the voice and the conversation is good, almost natural at times. It manages to be engaging most of the time. Some voices occasionally sound “robotic,” and the conversation can feel chaotic at times, especially in the Spanish version, where it is less clear who is interviewing whom and which voice belongs to the person with expertise on the topic. Sometimes a third voice appears, difficult to understand in the context of the previous interview.*
- *The scientific content is good, with useful metaphors to understand complex genetic concepts, both in English and Spanish. It provides an adequate summary of the article, covering the key topics and emphasizing fundamental points and learning aspects of the article being discussed. Occasionally, it can be confusing or repetitive, but most of the time, it contributes to and organizes the article’s information correctly. I would say that the quality of the English episode is higher than the Spanish one, both in content and form. It would be fantastic to clearly sepárate episodes for higher level listeners, as researchers or clinicians, and episodes for patients organizations or families.*
- *This tool has the potential to revolutionize how we address the knowledge and implementation gap in rapidly evolving fields, such as genetics or developmental epileptic encephalopathies. I see this as even more important in Spanish, where knowledge tends to reach the patient and medical Spanish-speaking community even later*.”

**Feedback 17:**

Alison Merket, Genetic Counseling Research Assistant at UTHealth, Houston, TX

“*Feedback about the podcasts, After listening to the SCN2A and SYNGAP1 podcasts:”*

- *“I think that these podcasts are an incredible tool that can help various audiences stay up to date with cutting edge research by breaking down complex topics into digestible and approachable pieces. In addition, they are extremely easy and enjoyable to listen to. I’m very impressed at the natural flow of the conversations between the two AI hosts making it feel like you are really sitting in the room with two relatable podcast hosts.*
- *In general, the hosts did a good job at guiding the conversation through different topics or probing questions, but I do believe that this is something that could be improved with our prompting. At the beginning of the podcast, it could be helpful to have the hosts give a brief summary or breakdown of the topics that will be covered in the podcast so the listener can be prepared before diving straight into a foreign topic. It could also be helpful to have a take-home message given at the end of the podcast so the listener can clearly understand the important points to take away.*
- *Overall, the podcast flowed well but there were some hiccups. For example, during the SCN2A podcast around time 14:53, the hosts bring the podcast to a close but then the podcast continues repeating a section that was previously played. This can easily be edited with editing software like iMovie but is a good example of why it would be important to have a human component reviewing the podcasts before uploading to a public platform.*
- *When posting these podcasts on YouTube, I think it would be a great opportunity to point the audience to any helpful resources. For example, if we are posting a podcast about SCN2A, we could list the FamilieSCN2A Foundation or any other resources that we know of which could be beneficial for families watching.”*

*Things to note:*

- *“After watching a couple of educational videos about NotebookLM and playing around with it a bit, there were a couple of interesting things to note: It seems like the only way to customize the podcast to your specifications is through the initial prompt that you feed it before generating the podcast. We can adjust this prompt and make it more specific, however, it only allows up to 500 characters which might be a limitation in some instances. For example, I tried to prompt AI to correct the pronunciation of SYNGAP1 throughout the podcast, but it only applied the correction to the first time SYNGAP1 was said. We could use more specific wording in the prompt about having the podcast correct the pronunciation every time, but we are limited to 500 characters. This is something we can continue working on or also look in to other AI tools that could tailor a podcast more specifically to our liking.”*

**Feedback 18:**

Victoria, Co-founder SHER Sociedad Hispana de Enfermedades Raras @SHER, Latin America Director SYNGAP1 Research Fund.

“*As Mike mentioned, at SRF, we firmly believe in the potential of podcasting and the immense value it can bring to our community. That's why we've created our Spanish podcast, Café Syngap1, where families can share their journeys to diagnosis. I am really excited about your Spanish podcast idea, as it offers a fantastic opportunity to provide updates in our rapidly evolving field. This initiative is a significant chance to communicate complex information in Spanish, particularly for caregivers and clinicians in Latin America who are in urgent need of information and updates about genetic encephalopathies. I did notice a few phrases that sounded a bit robotic, and there were moments where the host overlapped, but overall, the content effectively conveys important information and is shareable with our community and interested clinicians. I look forward to learning more about this project!”*

**Feedback 19:**

Anu Cherukara, Genetic Counseling Research Assistant, The University of Texas Health Science Center at Houston (UTHealth Houston)

*“Overall feedback summary: Initially hearing this reminded me of the DNA Today Podcast! It was fascinating to listen to an AI-driven podcast. These new tools and ideas will spread more awareness and empower one another to share information in the rare disease communities.  Personally, I find this helpful as someone who is preparing to become a genetic counselor. Also as a current GCA in the genetics research space, I can tell this will be a great way to keep professionals informed and applicable to their daily work”.*

**CACNA1A podcast:**

*“General feedback*

- *Timing of podcast: This podcast was 20 minutes long which is the perfect timing for those commuting or who has a fast-paced lifestyle, and busy schedule. An hour-long podcast is something people may not listen to in reality.*
- *Break in between podcast: I think there should be breaks in between a 30 minute podcast depending on the type of publication. I liked how there was a break in between the introduction and expanded on more topics introduced in the beginning of the podcast to the next part of the podcast.*
- *Introduction provided a brief overview of the topic, which was well-conversed.*
- *Research articles highlighting variability and delayed diagnoses in CACNA1A and treatment options for CACNA1A were summarized well and engaging to listen to.*
- *Defined genetic testing process and insights into variant classification and what a daily life looks like in affected individuals. Caregiver perspectives and experience were interesting and engaging to listen to.*

*Constructive Feedback*

- *Podcast platform channel uniqueness: (see DNA Today Podcast below)* [*https://www.youtube.com/channel/UCj5c3YSE2zMDaP4ATMrvKbg*](https://www.youtube.com/channel/UCj5c3YSE2zMDaP4ATMrvKbg)
- *Create a separate channel dedicated to podcasts we will be able to upload to similar to the example I showed above for organizational purposes and less clutter with other videos.*
- *Define purpose, title, citations etc: Educational podcasts for rare genetic disorders channel.*
- *CACNA1A pronunciation by first speaker was different from second speaker.*
- *There needs to be a way to credit this way of podcasting so people don’t get the wrong assumption that this was created by AI and not real people. This should be clear, so that misinterpretation and misunderstanding can be alleviated.*
- *I could have missed mistakes, but I believe this content is understandable and engaging to the general public audience.”*

***SLC6A1* podcast:**

“*General feedback*

- *Timing of podcast: About 16 minutes, which was easy to follow and a good average time for podcasts summarizing multiple publications/articles.*
- *The overall content was engaging and is valuable for raising awareness and creating community support.*

*Constructive feedback:*

- *Again in general just ethical concerns such as ensuring personal narratives from affected individuals and families are central to supporting material (publication).”*

**Feedback 20:**

Richard, neuroscience grad student intern and SRF volunteer.

*“I was honestly very impressed with the production quality. The podcast was set up in such a way that it is very conversational like Cafe Syngap which I liked. My only feedback for the Spanish version would be there are times in the audio clip where the pacing feels too fast which can lead to it sounding a bit robotic (e.g. 9:50-10:10). If there was a way to slow down the pacing for certain segments of the audio, that would be a big improvement.”*
