## Supplementary material for "Enhancing Rare Disease Education through AI-Driven Podcast Generation": Feedback 5

Outlook

### RE: MEDRXIV/2025/320972 -- Please Resubmit

**Desde** Alfred L. George <>
**Fecha** Lun 27-01-2025 15:54
**Para** Lal, Dennis <>; EDUARDO ESTEBAN PEREZ PALMA <>

Hi Dennis and Eduardo,

I give my consent to be acknowledged in this manuscript. My contributions were not sufficient for co-authorship.

Thanks

Al

Alfred L. George, Jr., M.D.
Professor and Chair
Department of Pharmacology
Northwestern University Feinberg School of Medicine
Searle 8-510
320 East Superior Street
Chicago, IL 60611

http://www.pharm.northwestern.edu/
https://www.youtube.com/watch?time_continue=6&v=BpU2nOtpuVM
https://epilepsy-channelopathy.org/

---

**From:** Lal, Dennis <>
**Sent:** Monday, January 27, 2025 11:27 AM
**To:** Alfred L. George <>;
**Subject:** FW: MEDRXIV/2025/320972 -- Please Resubmit

Hi Al,

Is it ok that we list you as author?

Otherwise, we need to consent you or remove your feedback. Eduardo will share with you the final version that you can make an informed decision.

Please see below.

Best
Dennis

--
Dennis Lal, PhD
Founding Director for the Center of Neurogenetics | UTHealth Houston, Texas, US
Associate Professor | McGovern Medical School, UTHealth Houston, Texas, US
Visiting Scientist | Broad Institute of Harvard and M.I.T., Cambridge MA, US
Consultant | Cleveland Clinic Neurological Institute, Ohio, US
Group Leader | University of Cologne, NRW, Germany
--
Science Twitter: @LalDennis


--
Check out webtools the research group has developed:
https://cacna1a-portal.broadinstitute.org
https://scn-portal.broadinstitute.org
https://grin-portal.broadinstitute.org
https://slc6a1-portal.broadinstitute.org/
https://simple-clinvar.broadinstitute.org
https://miscast.broadinstitute.org
https://per.broadinstitute.org
https://scn-viewer.broadinstitute.org/
https://ndd-cnv-portal.broadinstitute.org
https://vsranker.broadinstitute.org
https://scn1a-prediction-model.broadinstitute.org/
--
Here are our latest publications:
https://pubmed.ncbi.nlm.nih.gov/?term=Dennis+lal&show_snippets=off&sort=date&size=100
https://scholar.google.com/citations?user=FrNhW3oAAAAJ&hl=en

---

**From:** <>
**Date:** Monday, January 27, 2025 at 9:29 AM
**To:** <>
**Cc:** Lal, Dennis <>
**Subject:** MEDRXIV/2025/320972 -- Please Resubmit

External: Increase caution when handling links and attachments.

MS ID#: MEDRXIV/2025/320972
MS TITLE: Enhancing Rare Disease Education through AI-Driven Podcast Generation.

Dear Dr. Lal,

We are returning this new submission because it contains names of non-authors in the supplementary file "AI_Education_Podcast_Supplementary_File.docx". Please upload as supplementary information a consent document where these people authorize their names appearing in the manuscript as authors of the comments. Otherwise, please remove their names from the document if they are not listed as authors in the main manuscript.

You will find your submission in the "Papers Returned for Your Attention" section of your Author Area.

Kind regards,
The medRxiv team
